# Predictors of maternal and fetal outcomes of pregnant women with mitral stenosis

**DOI:** 10.1101/2022.08.04.22278407

**Authors:** Yoseph Gebremedhin, Senbeta Guteta, Binyam Melese, Elias Bekele

**Affiliations:** Debretabore comprehensive specialized hospital, Debretabore, Ethiopia; Addis Ababa University, School of medicine, Department of internal medicine, Cardiology unit, Addis Ababa, Ethiopia; Mojo general hospital, Mojo, Ethiopia

**Keywords:** Mitral stenosis, mitral valve area, maternal outcome, fetal outcome

## Abstract

**Background:** Valvular heart disease, in particular mitral stenosis, is associated with unfavorable pregnancy outcomes. Data on maternal and fetal outcomes of pregnant women with mitral stenosis in Ethiopia as well as in other sub-Saharan countries is scare. Therefore the aim of this study was to assess the predictors of maternal and fetal outcomes of pregnant women with mitral valve stenosis and to show the incidence of maternal and fetal complications as well.

**Method:** A retrospective cross section study was conducted. Medical records of pregnant women with mitral valve stenosis admitted at Tikur Anbessa Specialized Tertiary Referral hospital labor ward from January 2014 to December 2018 were reviewed. Data were entered and analyzed using SPSS version 25. Multivariate logistic regression analysis was used to evaluate associated factors. P-value of 0.05 was considered statistically significant.

**Result:** This study included 58 pregnant women with mitral stenosis. The mean age of the participants was 27(SD,±4.5) years. The mean mitral valve area among the participants was 1.1cm. About 79% of cases had mitral valve area of ≤1.5cm^2^ and 55.2% had valve area of ≤1cm^2^. Fifty one (87.9%) patients had moderate to severe pulmonary hypertension. The overall incidence of serious adverse pregnancy and peripartal outcome among the study participants was 44.8%. About 69% of patients had heart failure, from which 34.5% had NYHA class III and IV functional classes. Maternal death occurred in 4(6.9%) cases, whereas thromboembolism and ICU admission rates were 2(3.4%) and 3(5.2%) respectively. Neonatal complications including low birth weight, preterm delivery, abortions and still birth were 26.5%, 16%, 17.2% and 6.1% respectively. Mitral valve area was the main determinant factor for maternal and fetal outcomes of pregnant women with mitral stenosis (AOR=15, 2.50-90% CI, P=0.003).

**CONCLUSIONS:** The overall incidence of serious adverse maternal and fetal events in pregnant mothers with mitral stenosis was 44.5%. Mitral valve area was the main determinant factor for maternal and fetal outcomes of pregnant women with mitral stenosis (AOR=15, 2.50-90% CI, P=0.003)

## Introduction

Heart diseases are among the most common medical causes of maternal death during pregnancy. Maternal heart diseases comprise 0.2% to 3% of all pregnancies(1,2) and are responsible for 10% to 25% of maternal deaths(2). In developing countries including Ethiopia valvular heart diseases are frequently encountered cardiac lesions. It imposes significant morbidity and mortality during pregnancy, these includes congestive heart failure (CHF), arrhythmias, thromboembolic complications and maternal deaths. Stenotic valvular lesion and low cardiac output reduces blood flow to the uterus and placenta, hence results in fetal complications.

The prevalence of rheumatic heart disease is significantly high in developing world hence it is one of the commonly encountered cardiac lesion in pregnant women. Cardiac disease is an important cause of maternal death in developing nations. Rheumatic heart disease accounts for most of this mortality, and mitral stenosis being the commonest lesion(3). Maternal risks are increased with severe mitral valve stenosis(MVA<1cm^2^)(3). The overall mortality associated with mitral stenosis is reported to be 1%, but increases to 5% in NYHA class III and IV(4). Pregnancy is associated with new hemodynamic changes which occur during pregnancy, labor and in the postpartum period(5). These changes begin during the first five to eight weeks of pregnancy and reach their peak late in the second trimester(5). These physiologic changes includes: blood volume increases 43%(6), cardiac output increased by 40-43%(6), heart rate increased by 15-17 beat/minute(6) on top of this stroke volume become raised and vascular peripheral resistant gets decline(6). These changes return to pre-pregnancy baseline within 2 weeks following delivery(5). Pregnancy has a deleterious effect on stenotic valvular lesions(7). These physiologic changes which occur during pregnancy in the presence of stenotic mitral valve leads to elevated left atrial pressure and high trans-mitral pressure gradient which plays an important role in both maternal and fetal complications.

Most pregnant women who were symptom free become symptomatic during pregnancy due to the new hemodynamic changes and the development of symptom is an entry point for cardiac evaluation. This why, the diagnosis of mitral stenosis is made during pregnancy.

The aim of this study was to assess the predictors of maternal and fetal outcomes of pregnant women with mitral valve stenosis and to show the incidence of maternal and fetal complications as well. We believe that the finding of this study would show the magnitude of the problem along with the critical factors that lead to poor maternal and fetal outcomes.

## Methods and Materials

### Study Design, period and area

A 5year retrospective cross sectional study from January 2014 to December 2018 was conducted. The study was carried out at Tikur anbessa specialized hospital (TASH), which is located in Addis Ababa, Ethiopia. It is the largest teaching and referral hospital in the country, which has more than 700bed capacity.

### Study Population

Pregnant women with mitral valve stenosis

#### Inclusion Criteria

➣ All pregnant women with isolated mitral valve stenosis were included in this study.

#### Exclusion Criteria

➣ Pregnant women with valve lesions other than mitral stenosis, Prosthetics valves or other cardiac lesions were excluded from the study

#### Sample Size

➣ All pregnant mothers with mitral valve stenosis, who were admitted to TASH labor ward during the study period, from January 2014 to December 2018, were included.

#### Data Collection and Analysis

➣ Ethical clearance was obtained from Addis Ababa University, college of health science ethical committees, before proceeding to data collection and chart review. Afterwards medical records of all 58 pregnant women with mitral valve stenosis admitted at Tikur Anbessa Specialized Hospital (TASH) labor ward in the study period were reviewed. Predetermined clinical data including heart failure, arrhythmia, pulmonary edema, thromboembolism, intensive care unit admission, maternal death, abortion, preterm delivery, low birth weight, perinatal death and echocardiography parameters like mitral valve area & pulmonary hypertension were retrieved from the charts. Data were entered and analyzed using SPSS version 25. Descriptive statistics were used to analyze relevant socio-demographic and clinical characteristics. Multivariate logistic regression analysis was used to evaluate associated factors. P-value of 0.05 was considered statistically significant.

## Ethical Considerations

Ethical clearance was obtained from Addis Ababa University, college of health science ethical committees, before proceeding to data collection and chart review.

## Result

A total of 58 pregnant women with MS were included in this study. The mean age of the participants was 27 years with SD of 4.5. Most of the participants were from Addis Ababa. About 41.4% of the mothers were primiparous and the rest 58.6% were multiparous. The majority (89.7%) delivered by SVD and the rest 10.3% delivered by Cesarean section.

As shown in table 2&3, about 69% of the participants had heart failure. The NYHA functional class of the participants was 20.7% Class-I, 13.8% class-II, 15.5% class-III, and 19% class-IV. Pulmonary edema developed in 20.7% of cases.

**Table 1:** Socio-demographic and obstetrical profiles of the pregnant mothers with MS

| Variable |  | Number/percentage |
| --- | --- | --- |
| Residency | Addis Ababa | 39(67.2%) |
|  | Oromia | 14(24.1%) |
|  | Amhara | 3(5.2%) |
|  | SNNR | 2(3.4%) |
| Parity | Nulliparous | 24(41.4%) |
|  | Multiparous | 34(58.6%) |
| Mode of delivery | SVD | 52(89.7%) |
|  | C/S | 6(10.3%) |

**Table 2:** The NYHA class and prevalence of heart failure and pulmonary edema in pregnant mothers with MS

| Variable |  | Number/Percentage | Total |
| --- | --- | --- | --- |
| NYHA Class | Class-I | 12(30%) | 12(20.7%) |
|  | Class-II | 8(20%) | 8(13.8%) |
|  | Class-III | 9(22.5%) | 9(15.5%) |
|  | Class-IV | 11(27.5%) | 11(19%) |
| CHF | Yes | 40(69.0%) | 40(69.0%) |
|  | No | 18(31.0%) | 18(31.0%) |

**Table 3:**
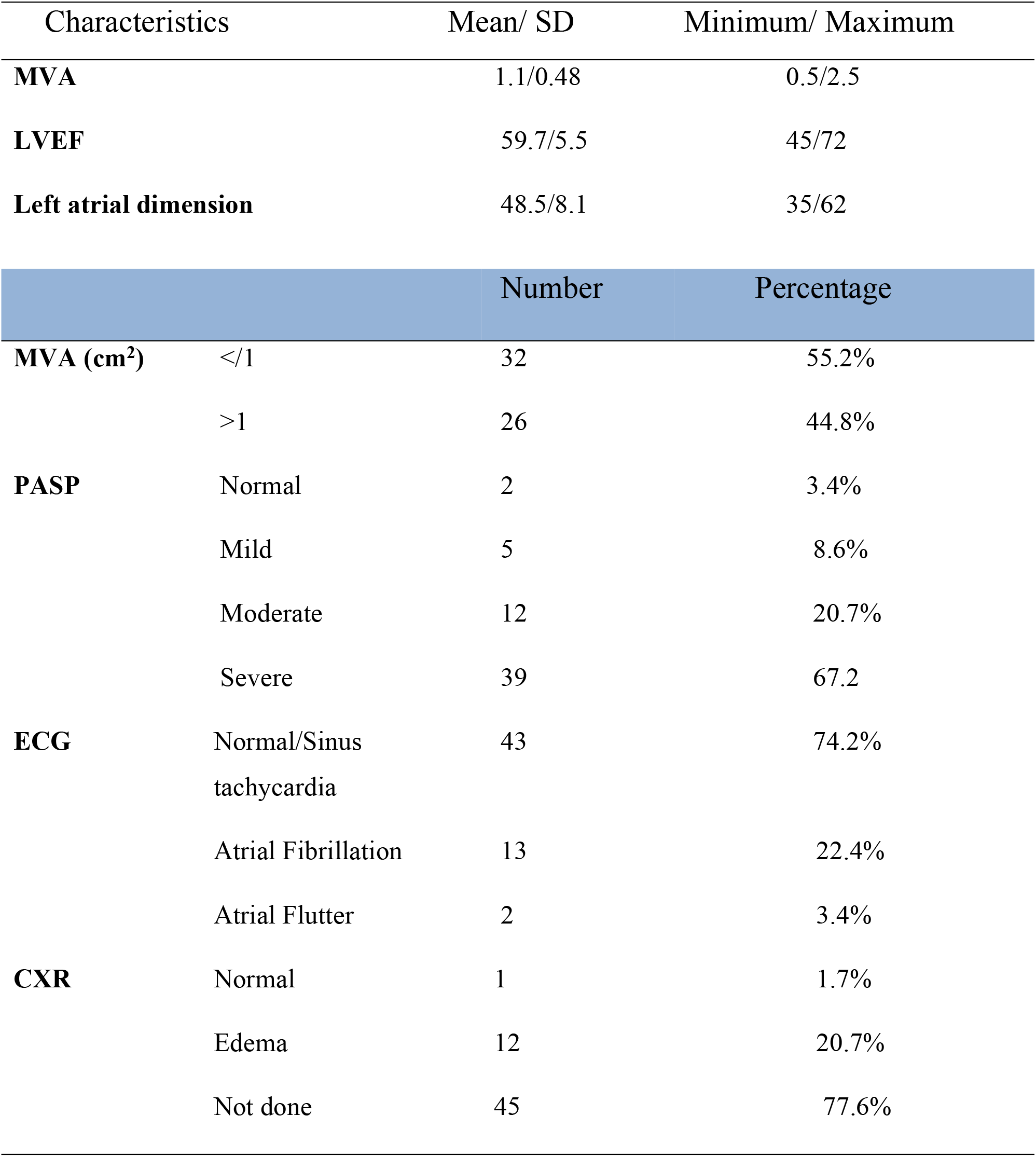
Echocardiography/ECG parameters and CXR results of the pregnant mothers with MS

The mean mitral valve area among the participants was 1.1cm (table 3). About 55.2% had severe mitral stenosis (MVA </1cm2). The pulmonary arterial systolic pressure was significantly elevated in most of the participants. About 67.2% of the participants had severe pulmonary hypertension, 20.7% had moderate pulmonary hypertension, and only 12% had mild or normal pulmonary artery systolic pressure. Among the abnormalities of cardiac rhythm, atrial fibrillation was present in 22.4% and atrial flutter in two patients.

The overall incidence of maternal and fetal unfavorable outcome among the study participants was 44.8 %(table 4). Four patients died, resulting in maternal death incidence of 6.9%, three patients required ICU admission, and two of them were among the cases who died. The other maternal complication was thromboembolism which occurred in two patients. Among the Neonatal complications, low birth weight occurred in 26.5%, abortion in 17.2%, 16% preterm deliveries and still birth in 6.1%.

**Table 4:** The type and proportion of unfavorable outcomes in pregnant mothers with MS

| Outcomes | Variables | Number /percentage |
| --- | --- | --- |
| **Unfavorable outcomes (overall)** | Yes | 26(44.8%) |
|  | No | 32(55.2%) |
| **Maternal complications** | Maternal Death | 4(6.9%) |
|  | ICU Admission | 3(5.2%) |
|  | Thromboembolism | 2(3.4%) |
| **Fetal complications** | Still Birth | 3(6.1%) |
|  | Abortion | 10(17.2%) |
|  | Pre term delivery | 8(16.0%) |
|  | Low birth weight | 13(26.5%) |

On multivariate analysis, as shown in table 5, the variables that were significantly associated with adverse pregnancy outcomes were PASP and MVA. However, on multivariate analysis (table 6) the only variable that remained statistically significant with adverse outcome was MVA </1cm2. The risk of adverse pregnancy outcomes in mothers who had MVA </1 cm2 was 15 times higher than mothers who had MVA >1cm2 (P-0.003, 95% CI 2.5-90)

**Table 5:**
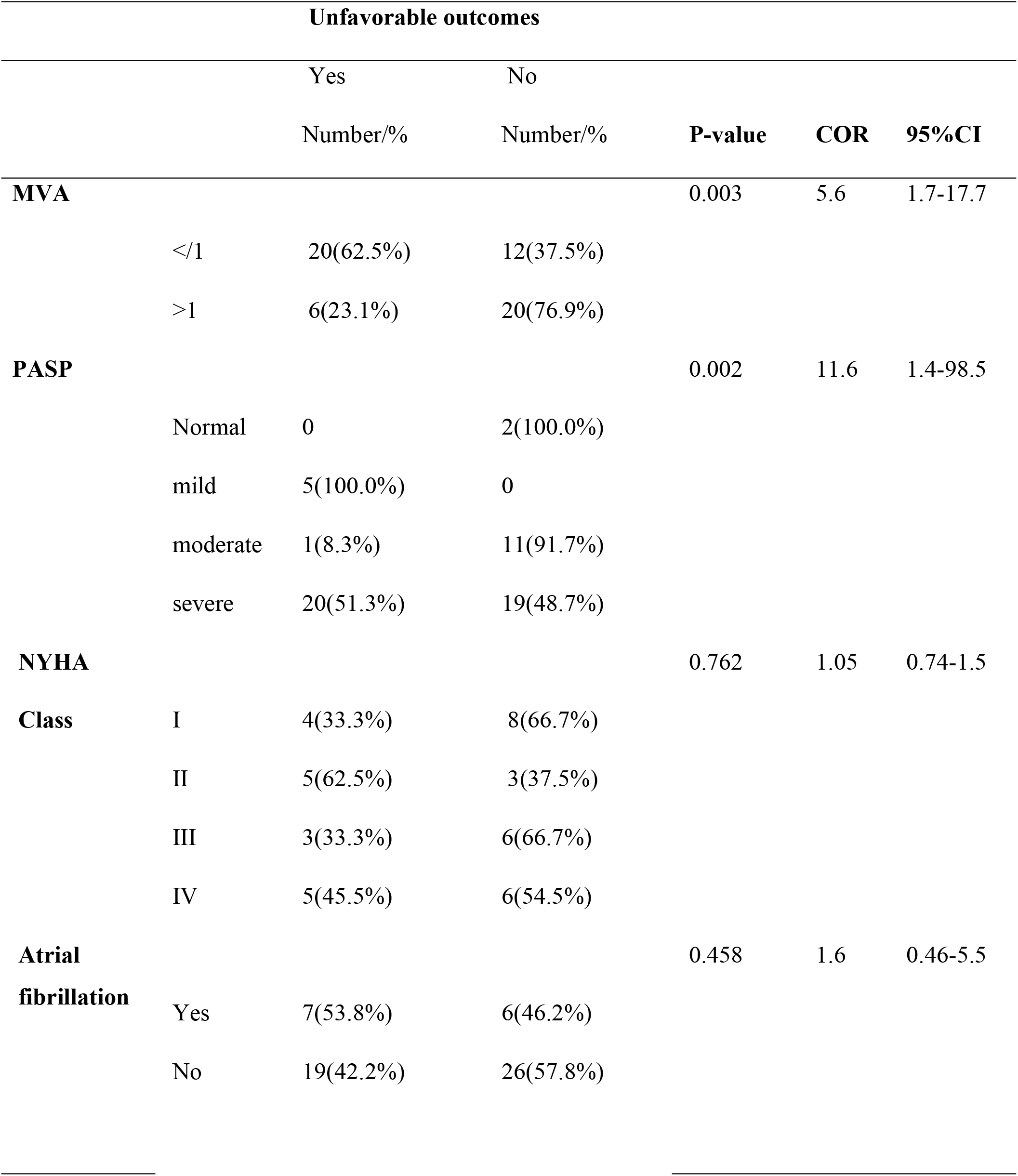
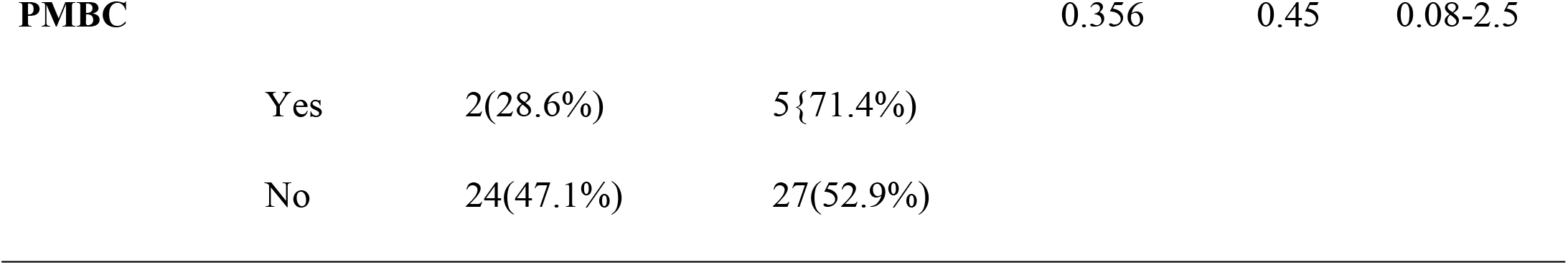
Association of Variables with unfavorable outcomes in pregnant mothers with MS

**Table 6:** Multivariate logistic regression of MVA and PASP with adverse pregnancy outcomes in MS

|  | **P-value** | **COR** | **AOR** | **95% C.I** |
| --- | --- | --- | --- | --- |
| **MVA </1cm<sup>2</sup>** | **0.003** | **5.6** | **15.00** | 2.50-90.02 |
| **PASP (severe)** | **0.095** | **11.6** | **2.43** | **0.86-6.87** |

**Table 7.** Comparison of maternal and fetal unfavorable outcomes in the current study with other studies done in patients with MS.

| Study | No | Severe MS | HF | Death | PTD | LBW | IUFD | Other |
| --- | --- | --- | --- | --- | --- | --- | --- | --- |
| California | 46 | 9 (19.5%) | 43% | 0% | 22% | 24% | 4% | ---- |
| ROPAC | 273 | 54(19.8%) | 23.1% | 1(1.9%) | 19.2% | 19.6% | 1.8% | Abortion<br>1.4% |
| Pakistan | 100 | 14(14%) | ---- | 2 | 21.8% | 27.7% | 9(9%) | ---- |
| SA | 80 | 7(9%) | 31% | 0% | 21.2% | 8% | 2.5% | ---- |
| Current study | 58 | 32(55.2%) | 69% | 4(6.8%) | 16% | 26.5% | 6.1% | Abortion<br>17.2%,TE<br>3.4% |

The incidence of unfavorable outcome was 2/7(28.6%) in mothers who had percutaneous mitral valvotomy whereas the remaining 5/7(71.4%) cases had successful pregnancy outcome.

When the probability of adverse event is plotted against MVA (figure 1), it progressively decreased as the MVA increases progressively.

**Figure 1:**
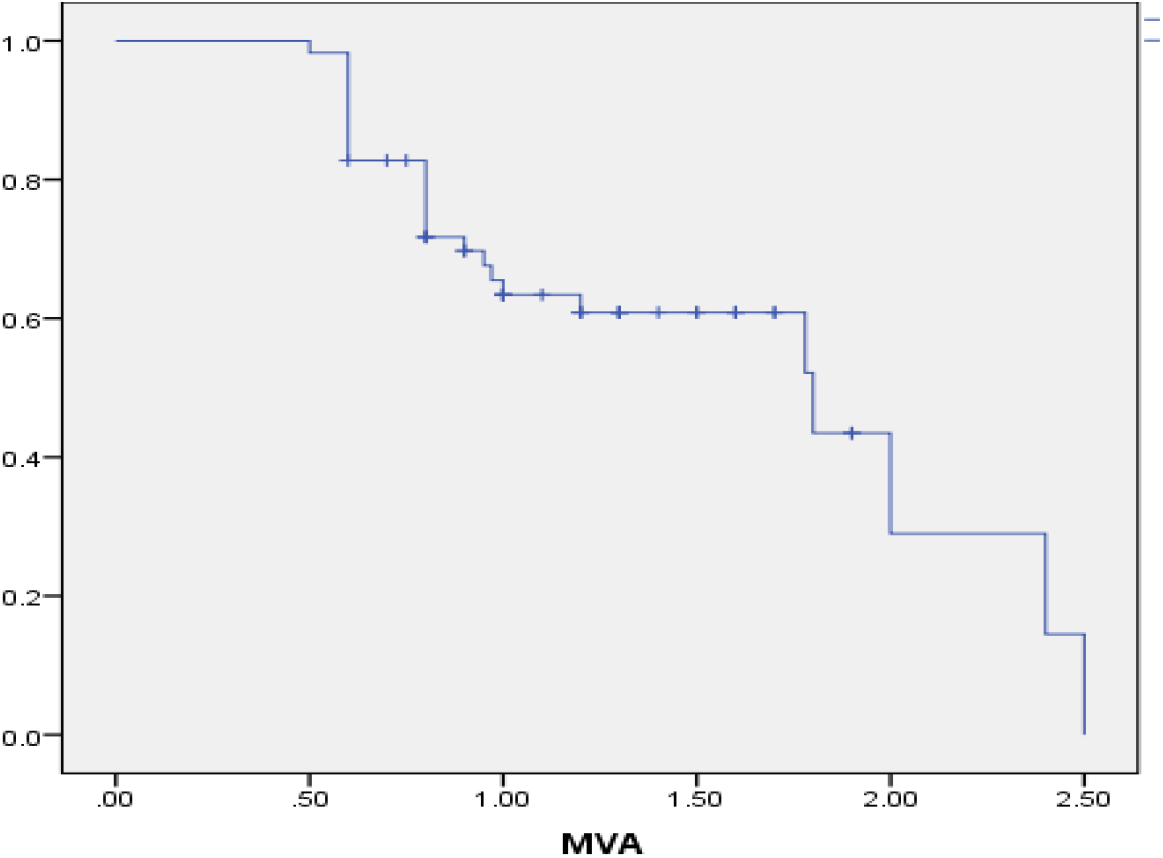
Probability of adverse outcomes plotted against MVA

The probability of adverse outcomes decreased progressively as the area of the mitral valve increases. The probability was highest in mothers with MVA of </1cm^2^

## DISCUSSION

A total of 58 pregnant women with MS were included in the study. The overall incidence of serious adverse pregnancy and peripartal outcome among the study participants was 44.8%. About 69% of patients developed heart failure, from this 34.5% had NYHA class III and IV functional classes. Maternal death occurred in 4(6.9%) cases, whereas thromboembolism and ICU admission rates were 2(3.4%) and 3(5.2%) respectively. Neonatal complications including low birth weight, abortions and still birth were 26.5%, 17.2% and 6.1% respectively.

The incidence of maternal death in the current study was 4/58 (6.8%). The incidence of maternal death in other studies was nil from studies in California and South America(8,9), 1/273 (0.37%) in registry of pregnancy and cardiac disease(10), and 2/100 (2%) in a study from Pakistan(11). Compared to these studies, the incidence of maternal mortality in our study is slightly higher. But unlike the other studies where most patients had mild to moderate MS, majority of the patients included in our study had severe MS. In addition, comparing mortality rate among these studies is difficult because the sample size in all of these studies is insufficient to determine mortality rate. However, from the finding of the current study and the reports of other studies mentioned above, it can be grossly assumed that maternal mortality is arguably rare in pregnant mothers with MS. The unique aspect of our study is that, most patients had the severe mitral stenosis, yet the incidence of death was not markedly different from other studies that included patients with higher number of mild & moderate MS.

The effect of optimal medical management in the outcomes of such patients is not also well studied. One study in south africa showed that pulmonary edema developed in about 31% of the patients, but all cases were successfully treated medically without any serious adverse maternal effects(9). In our study the overall rate of heart failure was 69%, from this 34% of cases had advanced NYHA class III and IV heart failure and 9 (20.7%) cases had radiologically confirmed pulmonary edema, all of these cases were successfully managed with medical therapies. In conclusion, even though the potential for maternal death for pregnant women with MS exists, its frequency is rare. Optimal medical management may decrease the risk of maternal death and other complications and should be applied as appropriate.

Among the fetal complications, LBW was the commonest complication in the current study which occurred in 26.5% of the deliveries. A comparable incidence of LBW was also reported from a study in California, Pakistan and Registry of Pregnancy and Cardiac Disease (ROPAC)(8,10,11). The incidence of IUFD in the current study was 6.1%. The incidence from other studies ranged from 1.8% to 9%(10,11). The incidence of abortion in our study was 17.2% which was markedly higher than the reports from other studies(10). This may be due to the cardiac profiles of the participants in our study. Majority of the patients in our study had severe MS. No other study had comparable proportion of patients with severe MS to that of our study. In conclusion, fetal complications are quite common in mothers with MS. The observed patterns of fetal complications indicate that decreased uteroplacental perfusion due to low cardiac output in MS is a likely explanation for these fetal complications. The biologic plausibility of this hypothesis can be inferred from the finding in our study and other studies which showed consistently strong association with fetal outcomes and the severity of the valve stenosis. These fetal complications can therefore be prevented by improving the cardiac output. Since MS is a mechanical problem, this can be achieved best by doing PBMC or MVR. Medical strategies to improve cardiac output such as treating atrial fibrillation, improving oxygenation, and other strategies as appropriate for the condition of the patient can also be beneficial in decreasing the risks of these fetal complications. Since fetal complications happened in all stage of pregnancy, medical treatment for the purpose of preventing such fetal complications should be started earlier in the course of pregnancy.

## Research Limitations

- Small sample size
- Single center, retrospective study

## CONCLUSION

The overall incidence of serious adverse maternal and fetal events in pregnant mothers with MS was 44.5%. The incidence of heart failure was 69% whereas maternal death occurred in 6.8% of cases. Mitral valve area was the main determinant of outcome in mothers with MS.

## Data Availability

All relevant data are within the manuscript and its Supporting Information files.

## What is already known on this topic

- Rheumatic heart disease is highly prevalent worldwide particularly in developing nations, hence it is one of the commonly encountered cardiac lesions in pregnant women.
- Cardiac disease is an important cause of maternal death in developing nations. Rheumatic heart disease accounts for most of this mortality, and mitral stenosis being the commonest lesion(3).
- However data on this topic is scarce in sub-Saharan countries where the problem is said to be high.

## What does this study adds

- The degree of mitral valve stenosis is the main determinant of unfavorable outcomes.
- The overall incidence of maternal and fetal unfavorable outcomes among pregnant women with mitral stenosis is 44.8 %.

## Authors’ contributions

Yoseph Gebremedhin, Binyam Melese and Elias Bekele conceived the study. Yoseph gebremedhin collected the data. Tegegn Molla and Yoseph Gebremedhin analyzed the data. Senbeta Guteta supervised all activities and commented on the manuscript. All authors confirmed the final version of the manuscript.

## Acknowledgement

I would like to express my deepest and heartfelt gratitude and sincere appreciation to Tikur Anbessa Specialized Tertiary Referral hospital staffs, especially Dr.Tegegn Molla who helped me a lot in data analysis.

I would like to thank Addis Ababa University, for granting me a chance to do this research.

